# The relationships between cochlear nerve health and AzBio sentence scores in quiet and noise in postlingually deafened adult cochlear implant users

**DOI:** 10.1101/2024.11.16.24317332

**Authors:** Zi Gao, Yi Yuan, Jacob J. Oleson, Christopher R. Mueller, Ian C. Bruce, René H. Gifford, Shuman He

## Abstract

**Objectives:** This study investigated the relationships between the cochlear nerve (CN) health and sentence-level speech perception outcomes measured in quiet and noise in postlingually deafened adult cochlear implant (CI) users.

**Design:** Study participants included 28 postlingually deafened adult CI users with a Cochlear® Nucleus™ device. For each participant, only one ear was tested. Neural health of the CN was assessed at three or four electrode locations across the electrode array using two parameters derived from results of the electrically evoked compound action potential (eCAP). One parameter was the phase locking value (PLV) which estimated neural synchrony in the CN. The other parameter was the sensitivity of the eCAP amplitude growth function (AGF) slope to changes in the interphase gap (IPG) of biphasic electrical pulses (i.e., the IPGE_slope_). Speech perception was tested using AzBio sentences in both quiet and a ten-talker babble background noise with +5 dB and +10 dB signal-to-noise ratios (SNR). IPGE_slope_ and PLV values were averaged across electrodes for each subject, both with and without weighting by the frequency importance function (FIF) of the AzBio sentences. Pearson and Spearman correlations were used to assess the pairwise relationships between the IPGE_slope_, the PLV, and age. Multiple linear regression models with AzBio score as the outcome and the PLV and the IPGE_slope_ as predictors were used to evaluate the associations between the three variables while controlling for age.

**Results:** The IPGE_slope_ and the PLV demonstrated different patterns with regards to their relationships with electrode location, age, and speech perception. The PLV, but not the IPGE_slope_, differed significantly across electrodes, where the apical electrodes had larger PLVs (better neural synchrony) than the basal electrodes. The IPGE_slope_, but not the PLV, was significantly correlated with participant’s age, where smaller IPGE_slope_ values (poorer spiral ganglion neuron density) were associated with more advanced age. The PLV, but not the IPGE_slope_, was significantly associated with AzBio scores in the +5 dB SNR condition, where larger PLVs predicted better speech perception. Neither the PLV nor the IPGE_slope_ was significantly associated with AzBio score in quiet or in the +10 dB SNR condition. The result patterns remained the same regardless of whether the mean values of the IPGE_slope_ and the PLV were weighted by the AzBio FIF. The result patterns generally did not change with fitting methods or input/output scales of the AGF slopes.

**Conclusions:** The IPGE_slope_ and the PLV quantify different aspects of CN health. The positive association between the PLV and AzBio scores in the +5 dB SNR condition suggests that neural synchrony is important for speech perception in adult CI users in challenging listening conditions with a relatively high noise level. The lack of association between age and the PLV indicates that reduced neural synchrony in the CN is unlikely the primary factor accounting for the greater deficits in understanding speech in noise observed in older CI users, as compared to middle-aged CI users.

## INTRODUCTION

The cochlear implant (CI), a prosthesis that partially restores hearing through stimulating the cochlear nerve (CN) via electrodes surgically implanted into the inner ear, is a standard treatment option for listeners with sensorineural hearing loss (for a review, see Zeng 2004).

Since electrical stimulation takes place at the auditory periphery, subsequent transmission of the signals by the CN is a prerequisite for the central auditory system’s ability to access and process the sound information. Therefore, it is believed that the neural health of the CN is crucial for the success of CI treatment (e.g., He et al. 2017; Zamaninezhad et al. 2023). The association between the CN health and hearing performance in CI users has been supported by post-mortem observations, where within-subject between-ear comparisons showed that the ear with a larger amount of spiral ganglion neurons (SGNs) consistently yielded a better word recognition performance (Seyyedi et al. 2014). However, due to the invasiveness of the histological procedures, direct examination of the CN is not feasible in living human CI users. Rather, non-invasive electrophysiological measures, such as the electrically evoked compound action potential (eCAP), have been developed to assess the CN health status of human CI users in research and clinical settings.

The eCAP is a near-field recorded, synchronized response of a population of CN fibers elicited by electrically stimulating a CI electrode, which has been used to evaluate neural encoding of electrical stimulation at the CN, such as spectral resolution (Won et al. 2014), neural adaptation (Hughes et al. 2012; He et al. 2023b), and amplitude modulation encoding (Tejani et al. 2017) in CI users. Morphologically, a typical eCAP waveform consists of a negative peak (N1) at around 0.2-0.4 ms after stimulus onset, followed by a positive peak (P2) at around 0.6-0.8 ms after stimulus onset (e.g., Brown et al. 1990). The amplitude of the eCAP waveform is defined as the difference in voltages between P2 and N1, and it increases with the stimulation level. The relationship between stimulation level and eCAP amplitude can be depicted using an amplitude growth function (AGF), also known as the input/output (I/O) function.

The slope of eCAP AGF has been shown to be associated with the density of SGNs in animal studies, where steeper slopes indicate higher SGN density in pharmaceutically deafened, implanted animals (Pfingst et al. 2015). Aligned with the animal results, shallower eCAP slopes were observed in pediatric CI users with cochlear nerve deficiency (CND) compared to those with normal-sized CNs (He et al. 2018). However, since the raw eCAP responses are susceptible to inter-patient and inter-electrode differences in non-neural factors (Brochier et al. 2021), such as electrode impedance and electrode-modiolus distance (Imsiecke et al. 2021), researchers have been seeking to overcome this drawback by using the differences between eCAP-derived measurements under various stimulation conditions to assess CN health. One particularly promising measurement is the sensitivity of the eCAP amplitude to changes in the interphase gap (IPG) of biphasic, electrical pulses (i.e., the IPG effects). Theoretically, non-neural factors should affect eCAP amplitude growth functions (AGFs) measured using different IPGs in a similar way. As a result, the effects of non-neural factors should be reduced or even eliminated when the differences between AGFs measured at different IPGs are calculated. Aligning with this theoretical possibility, research studies in both animal models and human CI users have demonstrated that the IPG effects are not affected by non-neural factors, such as electrode impedance (Schvartz-Leyzac & Pfingst 2016, 2018; Imsiecke et al. 2021) and electrode-modiolus distance (Schvartz-Leyzac et al. 2020b; Imsiecke et al. 2021).

In animal models, the IPG effects have been shown to be correlated with SGN survival.

Specifically, larger effects of IPG on eCAP amplitude (Prado-Guitierrez et al. 2006) and the AGF slope (Ramekers et al. 2014; Schvartz-Leyzac et al. 2019; Schvartz-Leyzac et al. 2020a) are associated with higher SGN density in guinea pigs. Consistent with these results measured in animal models, the IPG effect on the AGF slope (IPGE_slope_) has been shown to be positively correlated with sentence and consonant recognition (Schvartz-Leyzac & Pfingst 2018) and speech reception threshold (SRT; Zamaninezhad et al. 2023) in postlingually deafened adult CI users. More importantly, IPGE_slope_ was larger in children with normal-sized CNs than in children with CND (He et al. 2020; Yuan et al. 2022), a patient population with substantially less SGNs than other patient populations. Specifically, histological results of human temporal bone studies have shown that the mean SGN count in children with CND is 5,739 (range: 0-15,714), representing about 16.4% of the SGN count expected for the corresponding age, which is roughly 35,500 (Wright et al. 1986; Glueckert et al. 2010; Nelson & Hinojosa 2001; Haginomori et al. 2002; Chen et al. 2020; Da Costa Monsanto et al. 2022). In comparison, the mean SGN count in hearing impaired children with normal-sized CNs ranges from 14,301 to 20,523 (Miura et al. 2002). It should be noted that the axons of existing SGNs in children with CND are myelinated, with no apparent sign of degeneration (Glueckert et al. 2010; Wright et al. 1986).

Therefore, the results reported in He et al. (2020) and Yuan et al. (2022) provide a strong piece of evidence supporting the indicative value of IPGE_slope_ for SGN count. Nevertheless, the sensitivity of IPGE_slope_ as an index of SGN count in CI users cannot be determined due to the lack of direct and precise measurements of neural structure in living human listeners.

The IPG effect on stimulation level offset (IPGE_offset_) has been proposed as another candidate index of CN health. Based on computational modeling results, Brochier et al. (2021) proposed that the IPGE_offset_ outperformed the IPGE_slope_ in controlling for non-neural factors.

Consistent with this proposed relationship between IPGE_offset_ and CN health, Jahn and Arenberg (2020b) observed greater IPGE_offset_ in younger compared to older listeners. However, several empirical studies failed to demonstrate associations between the IPGE_offset_ and speech perception measurements in CI users with either full-length electrode array (Zamaninezhad et al. 2023) or short array to preserve residual low frequency acoustic hearing (Kim et al. 2010). Recent computational modeling studies on the IPG effects suggested that IPGE_slope_ and IPGE_offset_ might reflect different aspects of CN health. As noted in the discussion of Brochier et al. (2021), IPGE_offset_ likely reflects the properties and health status, rather than the number, of surviving SGNs. Aligned with this distinction, a three-dimensional model of the cochlea by Kipping et al. (2024) and Zhang et al. (2024) demonstrated correlations between IPGE_offset_ and CN degeneration, defined as reduced diameter in or entire loss of the peripheral axons. In contrast, a two-dimension model by Takanen et al. (2024) demonstrated that the IPGE_slope_ calculated as the numerical difference between the AGF slopes on a linear I/O scale is dependent on neural survival, and that non-neural factors such as electrode-neuron distance had little interference on the IPGE_slope_. Taken together, the existing evidence from animal, human and computational research are converging in suggesting that IPGE_slope_ is a promising indicator of CN density among the currently available eCAP measurements. As a result, IPGE_slope_ was used to assess neural survival of the CN in this study.

While having sufficient CN fibers responding to auditory input is a prerequisite for auditory perception, CN density alone does not guarantee good hearing functions in challenging listening environments. In theory, effective and accurate representation of sound signals that allows the listener to separate target signals from noise requires synchronous firing across neurons, which in turn depends on the health status of the CN, as demonstrated in animal and computational modeling studies (Kim et al. 2013; Heshmat et al. 2020). Neural desynchronization leads to a smeared representation of temporal cues, so that even though the ability to detect sound in quiet may be minimally affected, hearing performance in noise would degrade drastically. This scenario is exemplified by some listeners with auditory neuropathy spectrum disorder (ANSD), who have normal or relatively good behavioral audiometric thresholds and speech perception in quiet, but disproportionally impaired signal detection and speech perception in noise (Kraus et al. 2000; Zeng et al. 2005). The electrophysiological measures of patients with ANSD are characterized by a relatively normal cochlear microphonic and/or otoacoustic emission (OAE) response, and an abnormal or absent auditory brainstem response (ABR), which has been interpreted as a lack of synchrony across CN fibers despite a relatively normal hair cell function (e.g., Starr et al. 2008). The crucial role of neural synchrony in acoustic hearing has been further supported by the compound action potential (CAP) recorded in normal hearing (NH) listeners, where the level of neural synchrony, quantified with the phase locking value (PLV) of trial-by-trial CAP measurements, was found to be a strong predictor of recognition scores for speech in noise and time-compressed speech in quiet (Harris et al. 2021).

Due to the difference between acoustic and electrical hearing, the observations in listeners with NH or ANSD may not be readily generalizable to CI users. For many CI users, speech perception in noise is a challenging task despite excellent hearing performance in quiet (Zaltz et al. 2020). Histological observations of SGN dystrophy and demyelination in listeners with various hearing profiles (Nadol 1997; Wu et al. 2019) suggest that reduced neural synchrony could be an underlying cause of poor speech perception in noise in CI users.

However, this proposed relationship has rarely been evaluated, largely due to the lack of electrophysiological measures of neural synchrony in the CN. We recently developed a new method to quantify peripheral neural synchrony in CI users, where the PLV of trial-by-trial eCAP responses was used as an index to quantify the degree of neural synchrony in the responses generated by CN fibers across multiple electrical stimulations (He et al. 2024). We demonstrated that higher PLVs are associated with better temporal resolution and smaller effects of noise on word recognition in post-lingually deafened adult CI users, consistent with the hypothesized effect of neural synchrony on hearing performance in electrical hearing.

In summary, previous research has established both CN survival and neural synchrony as crucial factors contributing to hearing performance in CI users. However, considering that nerve damage can result in both lower neural density and poorer synchronization, as has been shown in computational models (Heshmat et al. 2020), little is known about whether the contributions of neural survival and synchrony to hearing performance are independent or overlapping.

Observations in NH listeners by Harris et al. (2021) suggest that neural engagement and synchrony are two separate dimensions that vary differently with changes in stimulus level. In human CI users, the number and synchrony of excited CN fibers have been modeled using retrospective deconvolution performed on intraoperative eCAP recordings, both significantly associated with postoperative speech recognition scores (Dong et al. 2023). However, the model was built upon assumptions about the shape of unitary response from CN fibers, which have not been directly validated in humans (Dong et al. 2020; Dong et al. 2023). While the IPGE_slope_ and the PLV in CI users have been measured postoperatively in separate experimental studies to assess their association with speech perception (Schvartz-Leyzac & Pfingst 2018; Zamaninezhad et al. 2023; He et al. 2024), it is unclear whether the biological underpinnings of these two indices are orthogonal and impact speech perception differently depending on listening conditions.

To address this critical knowledge gap, we measured the IPGE_slope_, the PLV and speech perception in the same group of postlingually deafened adult CI users and evaluated their relationships. Speech perception was measured using AzBio sentences (Spahr et al. 2012), a speech corpus consisting of multiple lists of everyday sentences with similar levels of difficulty. Based on previous studies on the effects of neural survival and neural synchrony on speech perception in listeners with various hearing profiles, we hypothesized that (1) the IPGE_slope_ and the PLV are two independent measures of CN health (Harris et al. 2021; Kraus et al. 2000); (2) the IPGE_slope_ is positively associated with speech perception in quiet (Zamaninezhad et al. 2023); (3) the PLV is positively associated with speech perception in noise (Zeng et al. 2005; Harris et al. 2021; He et al. 2024).

## MATERIALS AND METHODS

### Participants

Study participants included 28 postlingually deafened CI users (age range: 20.39-84.04 years, mean = 61.86 yrs, standard deviation *SD* = 14.78 yrs; 15 female, 13 male; nine bilateral CI users). Twenty-one of them also participated in our previous study on the development and validation of the eCAP PLV measurement (He et al. 2024), and their PLV data were reused in the current study. All participants were native speakers of American English and used a Cochlear® Nucleus™ device (Cochlear Ltd., New South Wales, Australia). All participants had a full 22-electrode insertion with their devices, as confirmed by postoperative computerized tomography scans. Only one ear was tested in each participant. For bilateral users, the tested ear was decided through randomization. With the exception of two participants (S21 and S25), none of the participants had any functional acoustic hearing in either ear. The lack of functional acoustic hearing was self-reported by the patients and was verified by pure tone audiometric thresholds of 70 dB HL or higher, averaged across octave frequencies between 250 Hz and 8,000 Hz. For the participants with functional hearing on the opposite side, an ear plug was used to minimize its potential contribution during the AzBio speech testing. Demographic information and hearing loss etiology of the participants are listed in Table 1. All participants provided written informed consent at their initial visit to the lab prior to data collection and were compensated for their time. The study was approved by the Biomedical Institutional Review Board at the corresponding author’s institution (No. 2017H0131).

**Table 1.** Demographic information of all participants. The participant IDs are not known to anyone outside the research group, including the participants themselves. M, male; F, female; L, left; R, right; SNHL, sensorineural hearing loss; CA, contour advance; DFNB8, deafness, autosomal recessive 8; DFNB10, deafness, autosomal recessive 10.

| Participant | Ear tested | Age range at testing (yrs) | Internal device and electrode array | Hearing loss etiology | Electrodes tested |
| --- | --- | --- | --- | --- | --- |
| S01 | L | 61-65 | CI 512 | Sudden SNHL | 3, 9, 14, 20 |
| S02 | L | 66-70 | CI 512 | Meniere's disease | 3, 9, 15, 18 |
| S03 | R | 66-70 | CI 24RE(CA) | Hereditary | 3, 9, 15, 21 |
| S04 | L | 36-40 | CI 24RE(CA) | Head Trauma | 3, 9, 15, 21 |
| S05 | R | 61-65 | CI 24RE(CA) | Unknown | 8, 12, 15, 18 |
| S06 | L | 51-55 | CI 532 | Unknown | 4, 9, 15, 21 |
| S07 | R | 61-65 | CI 522 | Head Trauma | 6, 9, 18, 20 |
| S08 | R | 36-40 | CI 24RE(CA) | Unknown | 3, 9, 15, 21 |
| S09 | R | 56-60 | CI 24RE(CA) | Unknown | 3, 9, 15, 21 |
| S10 | R | 66-70 | CI 532 | Unknown | 3, 9, 15, 21 |
| S11 | R | 66-70 | CI 532 | Unknown | 3, 9, 15, 21 |
| S12 | L | 76-80 | CI 422 | Unknown | 4, 9, 15, 21 |
| S13 | L | 61-65 | CI 632 | Unknown | 3, 9, 15, 21 |
| S14 | R | 66-70 | CI 24RE(CA) | Unknown | 3, 15, 21 |
| S15 | L | 66-70 | CI 532 | Vestibular<br>Schwannoma | 3, 9, 15, 21 |
| S16 | R | 81-85 | CI 532 | Unknown | 3, 7, 10, 17 |
| S17 | L | 56-60 | CI 532 | Unknown | 6, 9, 15, 21 |
| S18 | L | 76-80 | CI 622 | Unknown | 6, 9, 15, 21 |
| S19 | R | 66-70 | CI 632 | Head Trauma | 3, 9, 15, 21 |
| S20 | R | 81-85 | CI 632 | Unknown | 3, 9, 15, 21 |
| S21 | R | 51-55 | CI 632 | Unknown | 3, 9, 15, 21 |
| S22 | L | 56-60 | CI 632 | Unknown | 3, 15, 18 |
| S23 | L | 56-60 | CI 532 | Usher | 3, 9, 15, 21 |
| S24 | L | 76-80 | CI 632 | Unknown | 3, 9, 15, 21 |
| S25 | L | 36-40 | CI 612 | Vestibular<br>Schwannoma | 6, 15, 21 |
| S26 | L | 18-20 | CI 522 | DFNB8/10 | 3, 9, 15, 21 |
| S27 | L | 51-55 | CI 612 | Unknown | 5, 9, 15, 21 |
| S28 | R | 76-80 | CI 24RE(CA) | Unknown | 9, 12, 16 |

### Stimuli and Apparatus

For eCAP recordings, the stimulus was a charge-balanced, cathodic leading biphasic pulse with a pulse-phase duration of 25 µs. The IPG between the cathodic and anodic phases, as well as the presentation levels, varied across measurements, as detailed in the “Procedures” section. All eCAP recordings were performed using the neural response telemetry function implemented in the Custom Sound EP software v6.0 (Cochlear Ltd., New South Wales, Australia).

For speech perception tests, the stimuli were meaningful sentences (e.g., “The vacation was cancelled on account of weather.”) from the AzBio sentence corpus (Spahr et al. 2012), recorded by two female and two male native American English speakers. The sentences presented to each participant and for each condition were evenly distributed across the four speakers. The background noise was a ten-talker babble presented at two signal-to-noise ratios (SNRs): +10 dB and +5 dB. All stimuli were delivered via a loudspeaker (RadioEar Corporation, PA) placed 1 m in front of the participant at 0° azimuth in a sound-attenuated booth.

### Procedures

#### Testing Electrodes

The default testing sites for eCAP measurements were electrodes 3, 9, 15, and 21 (i.e., e3, e9, e15 and e21). These electrodes were selected to cover a wide range along the array with relatively equal numerical separations in between, while keeping the testing time reasonable. In the case of an open- or short-circuit or lacking a measurable eCAP at the maximum comfortable level due to high impedance at a default electrode, a nearby alternative electrode was tested. In rare cases when multiple neighboring electrodes are untestable, the sites were selected to cover a majority of the testable regions while keeping relatively equal separations in between. Four participants (S14, S22, S25, and S28) were tested at only three electrodes due to time constraints. The electrodes tested for each participant can be found in Table 1.

#### Behavioral C Level Measures

The maximum comfortable level (C level) for eCAP stimuli at each IPG level (7 µs and 42 µs) was determined via subjective rating using an ascending procedure. Prior to the measurement, participants were shown a visual scale of 1 (“barely audible”) to 10 (“very uncomfortable”) and were instructed to give a loudness rating using verbal responses or hand gestures following each stimulus presentation. Each presentation consisted of five pulses delivered at a probe rate of 15 Hz. The stimuli were first presented at a relatively low level and gradually increased in steps of 3-5 clinical levels (CLs) until a rating of “7” was reached, then in steps of 1-2 CLs until a rating of “8” was reached. The lowest level that corresponds to a rating of “8” (“maximal comfort”) was recorded as the behavioral C level.

#### eCAP Measures

The eCAP was measured using a two-pulse, forward-masking paradigm (Brown et al., 1990), where the masker pulse was always presented at 10 CLs higher than the probe pulse. The masker pulses were delivered at the testing electrode, and the eCAP responses were recorded two electrodes away from the testing electrode in the apical direction. There was an exception for electrode 21, which was recorded two electrodes away in the basal direction (i.e., electrode 19). The probe pulses were presented at a probe rate of 15 Hz with a masker-probe interval of 400 µs. The total number of trials in each stimulation sequence differed across measurements, as detailed below. Responses were recorded at a sampling rate of 20,492 Hz with a sampling delay of 122 µs, an amplifier gain of 50 dB, and a monopolar-coupled stimulation mode.

#### Measure of Neural Survival: The IPGE_slope_

In this study, the IPGE_slope_ was operationally defined as the numerical difference (in µV/dB, where dB was calculated in reference to 1 nC) between the AGF slopes with IPGs of 42 µs and 7 µs, and therefore, its measurement involved acquiring an AGF and calculating its slope at each IPG level for each participant. The IPG of 7 µs was chosen because it was the shortest IPG value available for eCAP recording in the Custom Sound EP software, and the IPG of 42 µs was selected to allow for comparisons with previous studies on IPG effects in CI users with a Cochlear® device (He et al. 2020; Yuan et al. 2022). For both IPG levels, the maximum presentation level of the stimuli was the behavioral C level measured with an IPG of 42 µs. To acquire AGFs, the eCAP measurement started at the maximum presentation level and decreased in steps of 1 CL for five steps, then in steps of 5 CUs until no peaks could be visually identified in the eCAP waveform, i.e., when the threshold is reached. Additional presentation levels in steps of 1 CL for five steps near and above the eCAP threshold were tested. At each presentation level, the eCAP waveform was acquired by averaging the raw responses to 50 pulses. The visual identification of eCAP peaks, or lack thereof, was performed by the experimenter at the time of testing and rechecked by an expert researcher offline.

The AGF slope at each IPG level was calculated using the window method developed by Skidmore et al. (2022), where linear regressions were performed on sliding windows along a resampled AGF, and the largest slope among all windows was regarded as the AGF slope. The window method was suitable for calculating the slope of AGFs with various shapes, both in animals and human listeners. For the results in human listeners, the correlation between AGF window slope and speech perception outperformed traditional fitting methods such as linear or sigmoid fitting. In the present study, we focused on the IPGE_slope_ results calculated using the window method in the linear-logarithmic I/O scale of µV/dB, following the recommendations by Skidmore et al. (2022). However, since linear fitting and the linear-linear I/O scale of µV/nC were commonly used in previous research on IPG effects (Schvartz-Leyzac & Pfingst 2018; Zamaninezhad et al. 2023), and that fitting method and I/O scale may affect how well eCAP measurements correlate with neural health and speech perception (Brochier et al. 2021; Skidmore et al. 2022), we also calculated the IPGE_slope_ on a linear-linear scale (µV/nC) with both linear and window fitting methods to evaluate the robustness of the results across fitting methods and I/O scales, briefly reported at the end of the Results section. All calculations were performed using MATLAB 2021b (MathWorks, MA).

#### Measure of Neural Synchrony: The PLV

For each participant and electrode, the PLV was derived from 400 eCAP trials measured using biphasic pulses with an IPG of 7 µs presented at the behavioral C level, using the method developed by He et al. (2024). The IPG of 7 µs was selected for PLV measurement because it is the default and most used IPG in the programming of Cochlear® CI devices. The PLV is a unitless value between 0 and 1, where 0 means that the phases were randomly distributed across trials, and 1 means that the phases were perfectly correlated. To calculate the PLV, the eCAP responses were re-referenced to the first sample of each trial and time-frequency decomposed at six linearly spaced frequencies (788.2, 1576.3, 2364.4, 3152.6, 3941.0, and 4729.2 Hz) and divided into six partially overlapped time frames with an onset-to-onset interval of 48.8 µs and a length of 1561.6 µs. At each frequency and within each time frame, the unit vectors representing the phases of the 400 individual trials were averaged, and the length of the averaged vector was taken as the time-frequency-specific PLV. The formula for calculating the PLV at the time *t* and the frequency *f* based on *N* individual trials is:

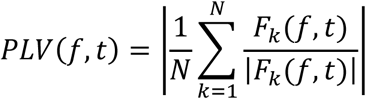

The PLV of the electrode was then obtained by averaging all time-frequency-specific PLVs calculated from the eCAP responses at that electrode. The time-frequency decomposition and the calculation of the PLV were performed using MATLAB R2021b and the newtimef.m function from EEGLAB v2022.1 (Delorme & Makeig 2004).

#### Measure of Speech Perception: AzBio Scores

Each participant was tested with AzBio sentences (Spahr et al. 2012) under three conditions: in quiet and in a ten-talker babble background noise with SNRs of +10 and +5 dB, respectively. The sentences were presented at 60 dB SPL in all conditions. The participants were tested unilaterally on the side with CI. Specifically, bilateral users were required to take off the CI on the opposite side. For the two participants with functional acoustic hearing in the opposite ear, an ear plug was used to minimize the contribution of acoustic hearing. For each participant and condition, a sentence list was randomly selected from Lists 1-8 of the AzBio corpus, each consisting of 20 sentences. For each participant, different word lists were used for different conditions. Participants were instructed to repeat back after each sentence and were encouraged to guess if they were unsure about what they heard. An experimenter recorded the number of words they correctly repeated in each sentence. The AzBio score was calculated as the number of words in the list correctly repeated by the participant, divided by the total number of words in the list. All words in the list, including prepositions, counted towards the score.

#### Averaging IPGE_slope_ and PLV Values across Electrodes

As demonstrated in He et al. (2023a), using eCAP results measured at single CI electrode locations to correlate with auditory perception outcomes in CI users can lead to inaccurate conclusions. Therefore, the values of the IPGE_slope_ and the PLV were averaged across all tested electrodes for each participant as an overall representation of CN health across the cochlea (He et al. 2023a). Both weighted and unweighted averages were calculated for both parameters. To calculate the weighted average, the results were weighted based on the frequency importance function (FIF) derived from AzBio scores under various spectral filtering conditions and SNRs in NH listeners (Lee & Mendel 2017). The FIF was fitted to a four-parameter Weibull function in SigmaPlot v15 (Grafiti LLC, CA). For each participant, the importance weight of each test electrode was calculated using the fitted Weibull function based on the electrode’s central frequency derived from the frequency-to-electrode table of the participant’s everyday programming map. The individual values of the empirically measured AzBio FIF and the fitted curve are shown in Figure 1. The unweighted average was calculated as the arithmetic means of the IPGE_slope_ and the PLV values across all tested electrodes for each participant.

**Figure 1.**
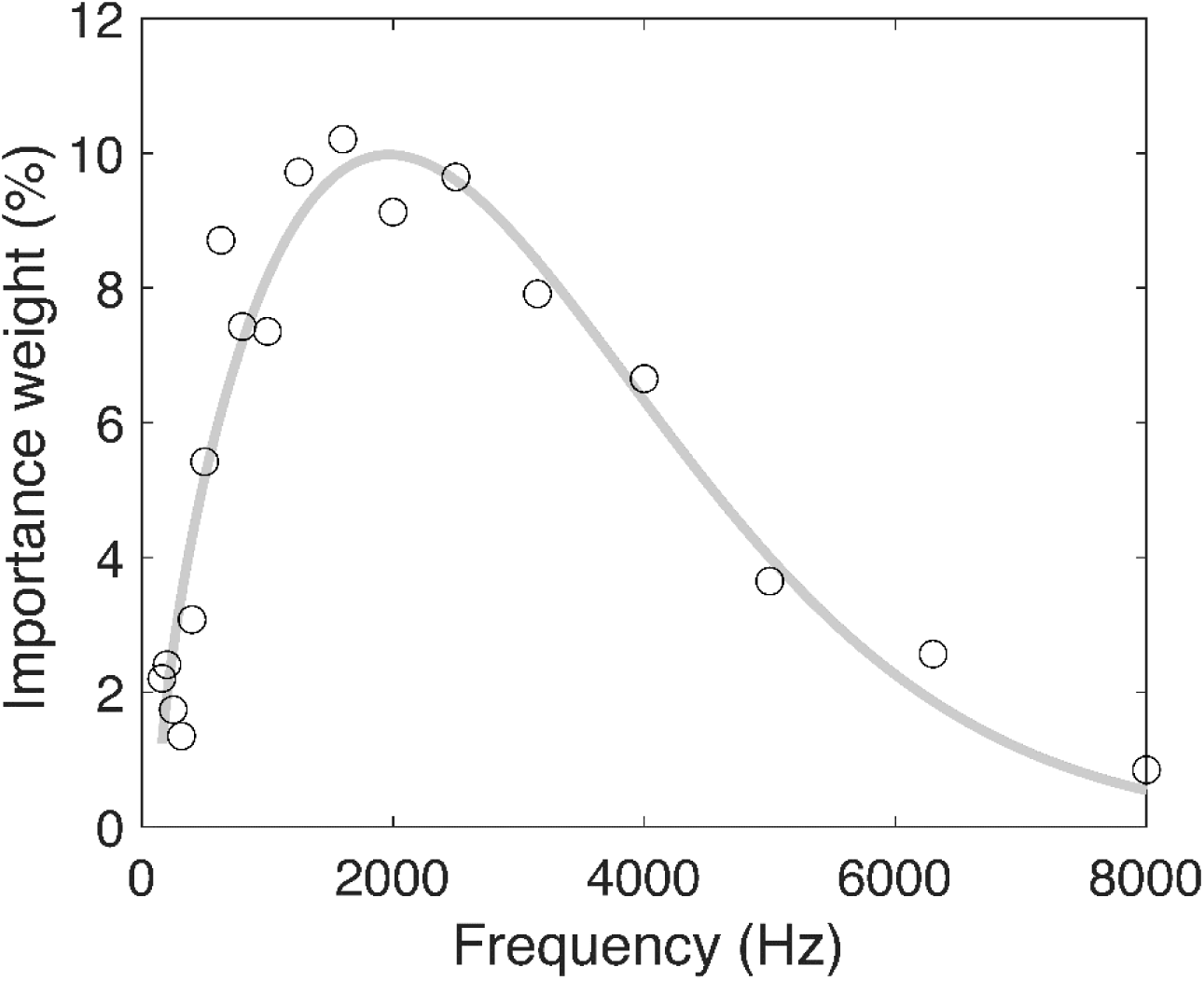
Individual AzBio FIF values (black circles) measured by Lee and Mendel (2017) and the fitted Weibull function (gray line)

### Data Analysis

The IPGE_slope_ analyses reported in the Results section are based on the AGF slopes obtained from the window fitting method in the unit of µV/dB, unless otherwise stated. The electrode-specific IPGE_slope_ and PLV values were compared across electrode locations using linear mixed-effect models. Pairwise comparisons between the electrodes were performed using the Tukey method for *p*-value adjustment. The IPGE_slope_ and PLV values were then averaged across the tested electrodes, weighted by the AzBio FIF, as representations of the overall CN neural health. The pairwise relationships between the IPGE_slope_, the PLV, and age at testing were assessed using either Pearson or Spearman correlation tests for variable pairs depending on the results of the Shapiro-Wilk normality test. Multiple linear regressions with AzBio score as the outcome and the IPGE_slope_ and the PLV as predictors were performed to evaluate the associations among the three variables under each testing condition (quiet, +10 dB, and +5 dB SNR).

Participant’s age was added to the regression models as a covariate to control for the potential effects of advanced age on speech perception and/or eCAP measurements, as have been demonstrated in CI users (Roberts et al. 2013; Sladen & Zappler 2015; Xie et al. 2019; Jahn & Arenberg 2020a). If the residuals were not approximately normally distributed, the outcome variables were transformed with appropriate methods to ensure that the normal residual assumption of linear regression is met. Correlation tests were performed in JASP v0.18.3 (JASP Team 2024), and the regressions were performed in R v4.4.1 (R Core Team 2024), with the lme4 (Bates et al. 2015), emmeans (Lenth 2024), and lmerTest (Kuznetsova et al. 2017) packages used for the linear mixed-effect models.

## RESULTS

The individual IPGE_slope_ and PLV values measured at each electrode are shown in Figure 2, with the range, mean, and standard deviation (SD) listed in Table 2. To assess the potential differences in the two measurements across electrodes, two linear mixed-effect regressions were performed with the IPGE_slope_ and the PLV as the outcome variables, the electrode location as the fixed effect, and participant as the random effect. The four categories of electrode locations corresponded to and were named after the four default testing locations: e3, e9, e15, and e21. If other electrodes were measured in lieu of the default electrodes, they were assigned to one of the categories based on their locations relative to the other electrodes tested in the same participant. For example, among the four electrodes tested in participant S05, e8 was the most basal electrode and therefore categorized as “e3” in the regression model. The category “e3” was used as the reference level in the regressions. The degrees of freedom were estimated using the Satterthwaite’s method. The results reveal an overall effect for PLV across electrodes (*F_(3, 77.4)_* = 5.44, *p* = .002). Focusing on the pairwise comparisons, only two comparisons showed a statistically significant difference in PLVs which were e3 compared with the PLVs measured at e15 (*t_(77.2)_* = 3.47, *p* = .005) and e21 (*t_(77.2)_* = 3.27, *p* = .009). The other comparisons were not significantly different which were e3 with e9 (*t_(77.7)_* = 1.28, *p* = .578), e9 with e15 (*t*_(77.5)_ = 2.09, *p* =.167), e9 with e21 (*t*_(77.5)_ = 1.89, *p* =.240), and e15 with e21 (*t*_(77.0)_ = 0.20, *p* =.997). For the analysis of electrode location effect on the IPGE_slope_, two data points (S08 e9 and S26 e21) were excluded from the analysis because their IPGE_slope_ values were more than three *SDs* away from the mean of the corresponding electrode location. The IPGE_slope_ did not significantly differ across electrodes (*F_(3, 75.2)_* = 2.22, *p* = .092). In the subsequent sections, the IPGE_slope_ and the PLV refer to the weighted averages of the corresponding values across electrodes for individual participants unless otherwise stated. It is worth noting that no outliers were removed in the subsequent analysis, as all PLV and IPGE_slope_ values fell within three *SDs* from the mean when averaged across all tested electrodes of each participant.

**Figure 2.**
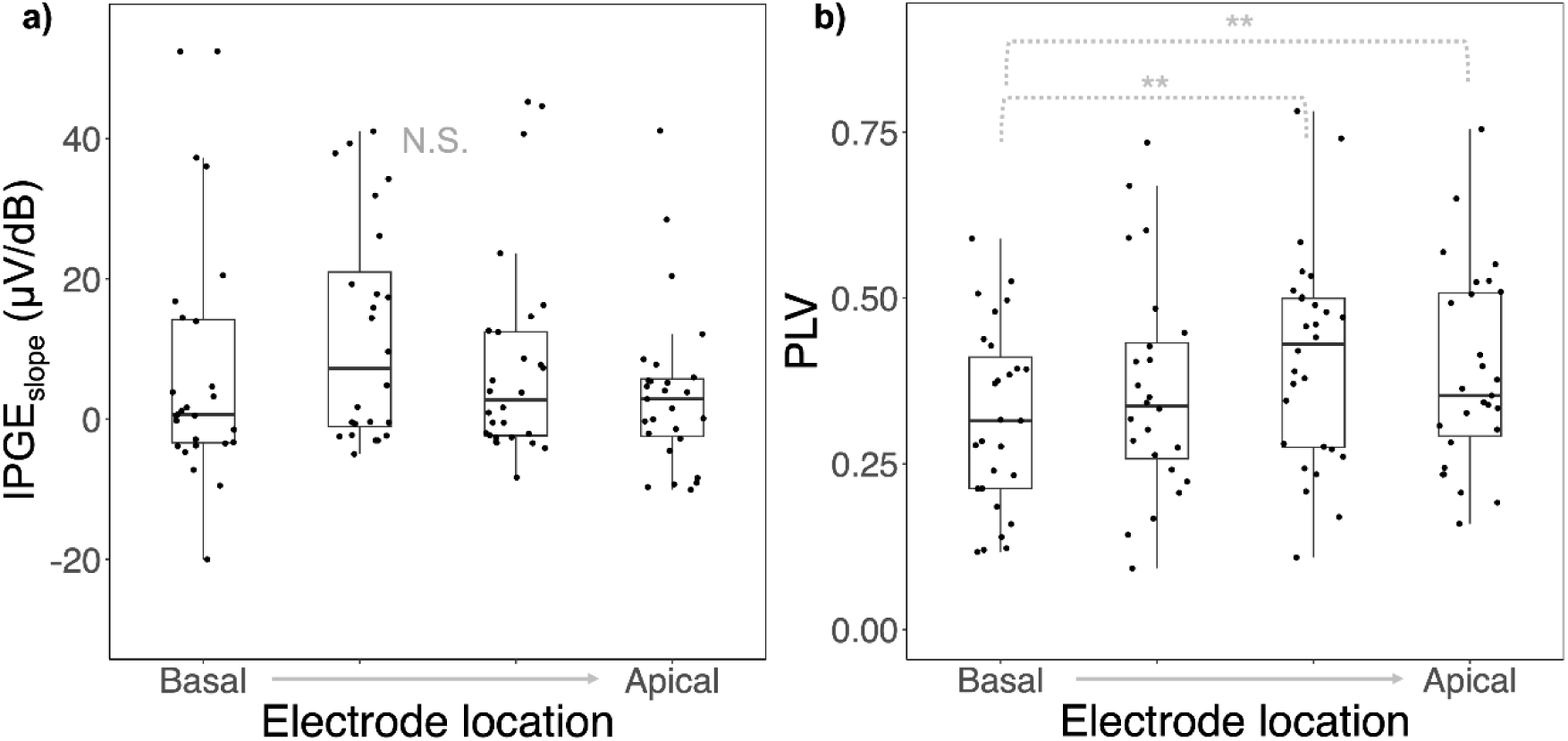
Individual values of (a) the IPGE_slope_ and (b) the PLV by electrode location. The four categories from basal to apical locations corresponded to, and were named after, the default electrodes e3, e9, e15, and e21, respectively. Values measured at non-default electrodes were categorized based on their locations relative to the other electrodes tested in the same participant. Boxes show the range between the first and the third quartile of the data values. The horizontal bars inside the boxes represent the median. The vertical whiskers show the range of values that are within 1.5 interquartile range (IQR) from the boxes. Asterisks denote pairwise comparisons with significant differences (*p* < .05). N.S., no significant differences.

**Table 2.**
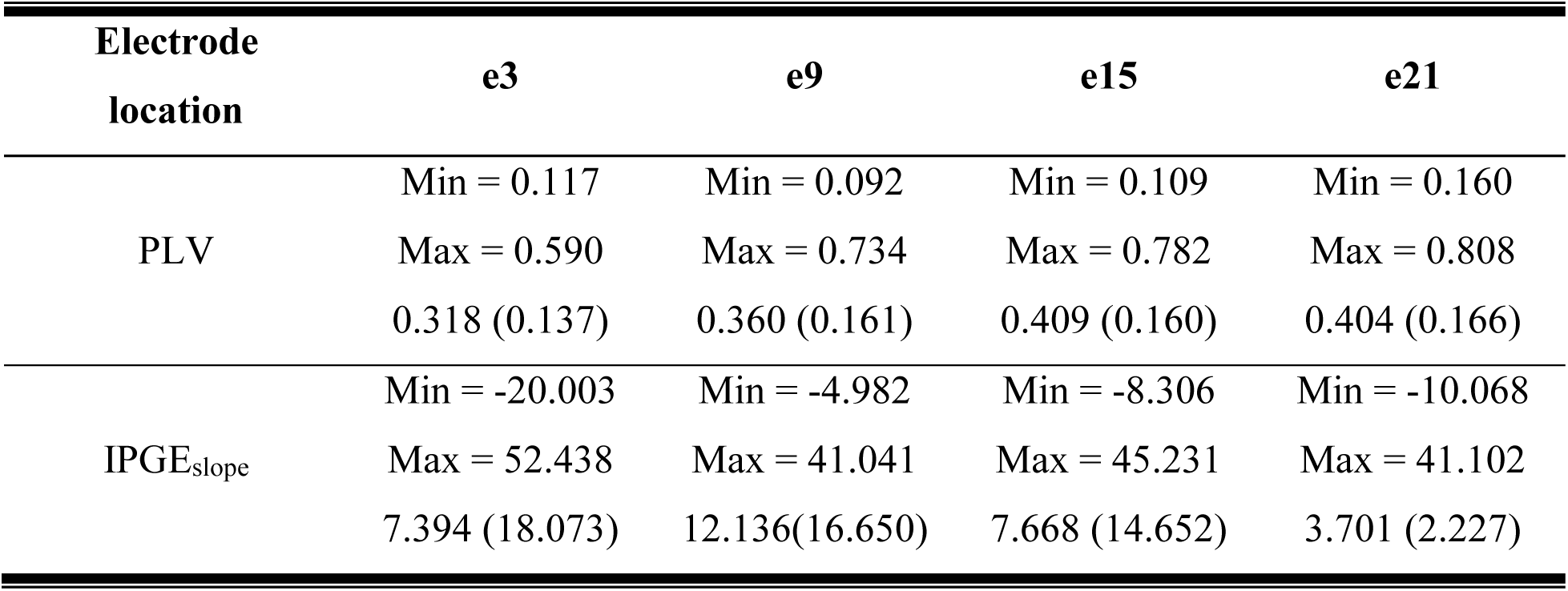
Descriptive statistics of the IPGE_slope_ and the PLV values measured at different electrode locations. The IPGE_slope_ values were calculated from AGF slopes in µV/dB fitted using the window method. Two electrodes were identified as outliers for IPGE_slope_ (more than three *SDs* away from the mean) and therefore excluded from the descriptive statistics. Values in each cell are listed in the format of “min, max, mean (*SD*)”.

| Electrode<br>location | e3 | e9 | e15 | e21 |
| --- | --- | --- | --- | --- |
| PLV | Min = 0.117 | Min = 0.092 | Min = 0.109 | Min = 0.160 |
|  | Max = 0.590 | Max = 0.734 | Max = 0.782 | Max = 0.808 |
|  | 0.318 (0.137) | 0.360 (0.161) | 0.409 (0.160) | 0.404 (0.166) |
| IPGE<sub>slope</sub> | Min = -20.003 | Min = -4.982 | Min = -8.306 | Min = -10.068 |
|  | Max = 52.438 | Max = 41.041 | Max = 45.231 | Max = 41.102 |
|  | 7.394 (18.073) | 12.136(16.650) | 7.668 (14.652) | 3.701 (2.227) |

### Correlations between the IPGE_slope_, the PLV, and Age

The eCAP AGFs acquired at different IPG durations from one example participant (S06) are shown in Figure 3. Time-frequency specific PLV values of the same participant are illustrated in Figure 4. Figure 5 shows the standardized values of IPGE_slope_ and PLV of individual participants (panel a), along with their age (panels b-c). All values were within three *SDs* from the mean. Spearman correlation tests showed that there is a non-significant trend that the IPGE_slope_ and the PLV were positively correlated (*rho_(26)_* = 0.340, *p* = .077). Although not a focus of this study, we assessed the relationships between age and the two CN health measures. Spearman correlation test results revealed a significant negative correlation between the IPGE_slope_ and age (*rho_(26)_* = -0.583, *p* = .001). Pearson correlation test results showed that PLV was not significantly correlated with age (*r_(26)_* = -0.068, *p* = .730).

**Figure 3.**
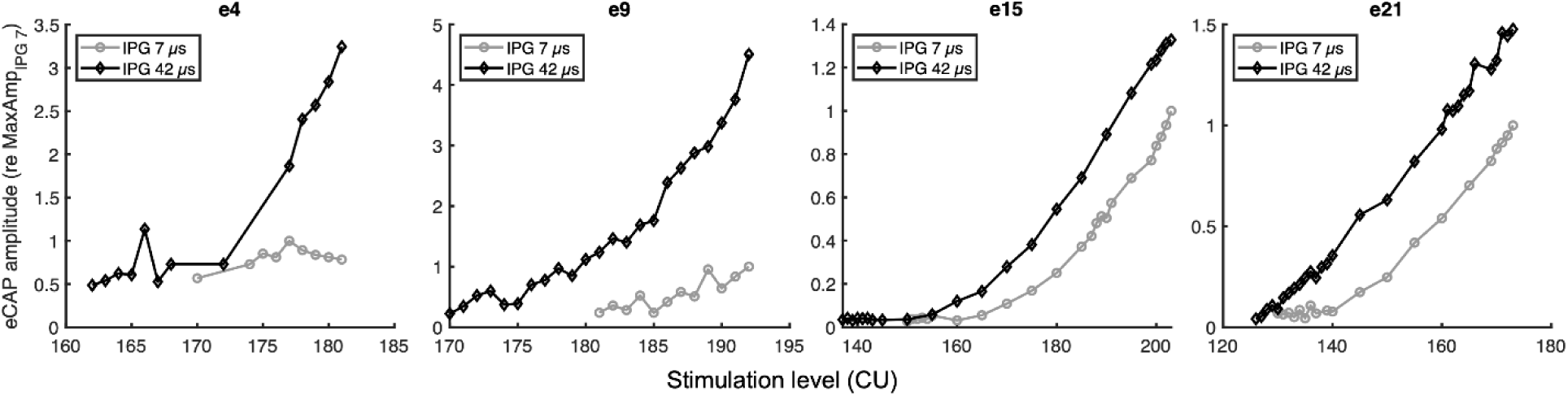
The AGFs measured with IPGs of 7 µs and 42 µs in one participant (S06), where the maximum stimulation level was set to the C level measured with a 42-µs IPG at each electrode. The amplitudes were normalized by dividing the amplitude of each trial by the maximum amplitude among the trials with a 7-µs IPG. Please note that the ranges of the axes are different across panels due to large variabilities in eCAP thresholds, C-levels, and amplitudes across electrodes.

**Figure 4.**
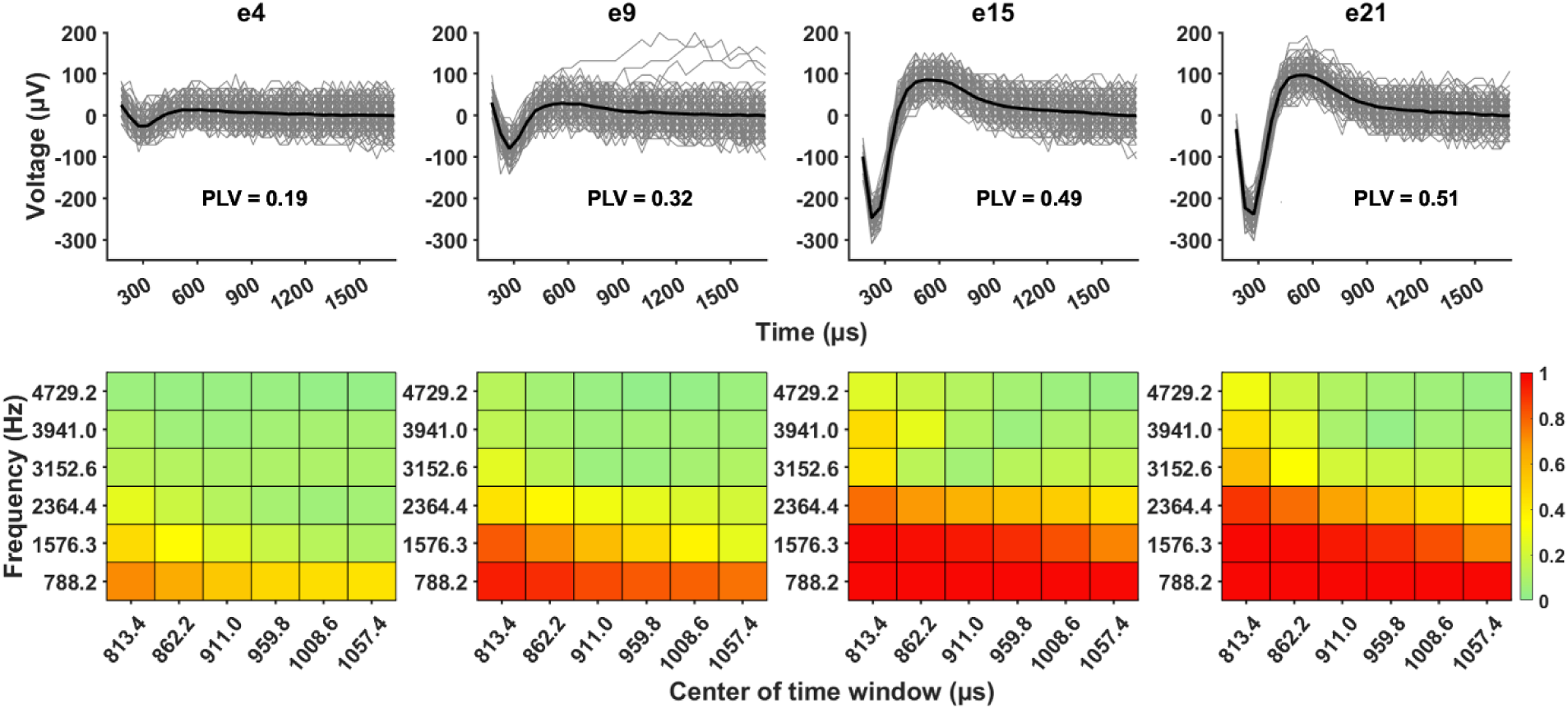
PLV values measured in one participant (S06). The time-frequency specific PLV values are shown in the heatmaps in the lower panels. The overall PLV values of individual electrodes are listed in the upper panels. Bolded black lines represent eCAP responses averaged across 400 trials. Responses in individual trials are plotted with gray lines.

**Figure 5.**
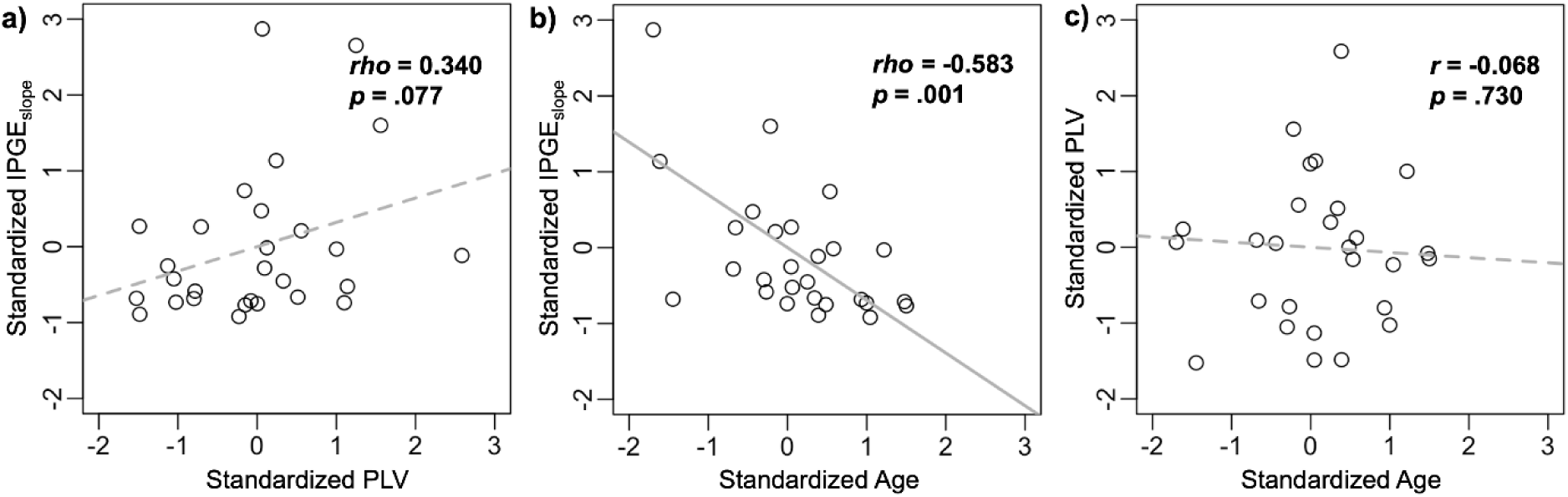
Correlations between (a) the IPGE_slope_ and the PLV, (b) the IPGE_slope_ and age, and (c) the PLV and age. The axes were standardized by converting into z-scores to improve comparability across panels. Each symbol indicates the result measured in one participant. The results of correlation tests are shown in each panel.

### The Associations among the IPGE_slope_, the PLV and Speech Perception Scores

The relationships between AzBio results in quiet and noise and eCAP measurements are illustrated in Figure 6. The AzBio scores in quiet were rank transformed (i.e., the lowest and highest scores were transformed into 1 and 28, respectively) due to non-normally distributed residuals when the raw scores were used in the linear regression model (not reported here). The results of linear regressions “AzBio ∼ PLV + IPGE_slope_ + age” revealed significant associations between the PLV and AzBio scores in the noise +5 dB SNR condition (*t_(24)_* = 2.36, *p* = .027) after adjusting for IPGE_slope_ and age, where larger PLVs (better neural synchrony) are associated with higher AzBio scores, but not in the noise +10 dB SNR (*t_(24)_* = 1.61, *p* = .121) or the quiet condition (*t_(24)_* = 0.71, *p* = .485). No significant association between the IPGE_slope_ and AzBio scores was observed in any testing conditions (*p* > .10 in all cases) after adjusting for PLV and age. Detailed results of the linear regression models are available in Table 3. It is worth noting that rank-transformation of the AzBio scores in quiet did not change the pattern or statistical significance of the results.

**Figure 6.**
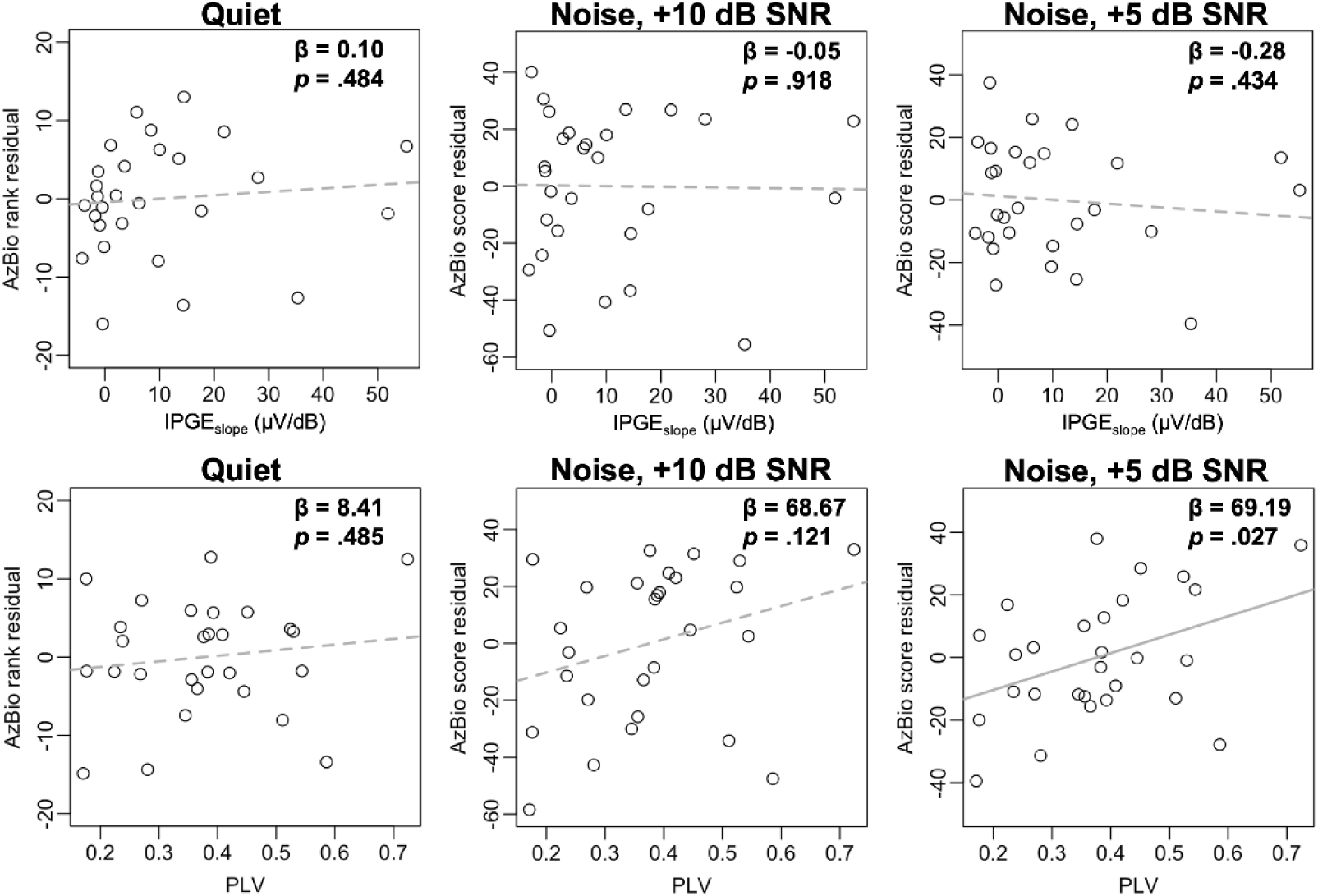
The residuals of AzBio results measured in quiet and in noise with +10 and +5 dB SNRs calculated using reduced linear regression models, plotted against eCAP measurements. In the top panels, vertical axes represent the residuals of “AzBio ∼ PLV + age” to highlight the relationships between IPGE_slope_ and AzBio results. In the bottom panels, vertical axes represent the residuals of “AzBio ∼ IPGE_slope_ + age” to highlight the relationships between PLV and AzBio results. The unstandardized beta and *p*-values were obtained from the full model “AzBio ∼ IPGE_slope_ + PLV + age” (Table 3) for the corresponding eCAP measurements shown in the horizontal axis of each panel. Each symbol represents the AzBio result measured in one participant.

**Table 3.** Results of linear models examining the relationships between the IPGE_slope_, the PLV, and AzBio scores measured in quiet and in two noise conditions. The IPGE_slope_ values were calculated from AGF slopes in µV/dB fitted using the window method. The AzBio scores in the quiet condition were rank transformed to meet the normal residual assumption of linear regression.

| Listening condition | Predictor | Unstandardized $\beta$ (SE) | $t$ | $p$ | Multiple $R^2$ |
| --- | --- | --- | --- | --- | --- |
| Quiet | IPGE<sub>slope</sub> | 0.1003 (0.1411) | 0.711 | .4839 | 0.2312 |
|  | PLV | 8.4126 (11.8582) | 0.709 | .4849 |  |
|  | age | -0.1561(0.1421) | -1.098 | .2829 |  |
| Noise, +10 dB SNR | IPGE<sub>slope</sub> | -0.0525(0.5075) | -0.104 | .9184 | 0.1555 |
|  | PLV | 68.6727 (42.6512) | 1.610 | .1205 |  |
|  | age | -0.4376 (0.5111) | -0.856 | .4004 |  |
| Noise, +5 dB SNR | IPGE<sub>slope</sub> | -0.2777 (0.3490) | -0.796 | .4340 | 0.3161 |
|  | PLV | 69.1906 (29.3348) | 2.359 | .0268* |  |
|  | age | -0.7400 (0.3515) | -2.105 | .0459* |  |

### Electrode Weighting by AzBio FIF

To qualitatively evaluate whether weighting the IPGE_slope_ and the PLV by the AzBio FIF modifies their relationships with AzBio scores, in a separate set of linear regressions, we used unweighted averages of the IPGE_slope_ and the PLV in lieu of their weighted counterparts. Overall, the result patterns and statistical significance remained the same in the unweighted version of the linear regressions, where the relationships between the PLV and AzBio scores were statistically significant in the noise +5 dB SNR condition (*t_(24)_* = 2.47, *p* = .021) but not in the noise +10 dB SNR (*t_(24)_* = 1.74, *p* = .094) or the quiet condition (*t_(24)_* = 0.91, *p* = .374), and no significant relationship between the IPGE_slope_ and AzBio scores was observed in any of the tested conditions (*p* > .10 in all cases).

### Comparisons across AGF Fitting Methods and I/O Scales

To evaluate the potential effects of AGF fitting methods and I/O scales on the results, we recalculated the IPGE_slope_ based on AGF slope used linear fitting and the window method, both in the linear-linear I/O scale of µV/nC. The statistical tests described above were then reperformed using each version of the IPGE_slope_ values. The result patterns and statistical significance stayed largely the same across fitting methods and I/O scale, with a couple exceptions. The non-significant trend of positive correlation between the IPGE_slope_ and the PLV was shared when the IPGE_slope_ values were calculated using window fitting in µV/dB (*rho_(26)_* = 0.340, *p* = .077) and linear fitting in µV/nC (*rho_(26)_* = 0.352, *p* = .067), but became a significant positive correlation when IPGE_slope_ was calculated using window fitting in µV/nC (*rho_(26)_* = 0.490, *p* = .009). The non-significant association between the PLV and the AzBio scores in noise with +10 dB SNR was shared when the IPGE_slope_ values were calculated using window fitting in µV/dB (*t_(24)_* = 1.61, *p* = .120) and in µV/nC (*t_(24)_* = 1.62, *p* = .119), but became a significant positive association when IPGE_slope_ was calculated using linear fitting in µV/nC (*t_(24)_* = 2.35, *p* = .028).

## DISCUSSION

This study assessed the relationships between two CN health measures, the IPGE_slope_ and the PLV, and evaluated their contributions to speech perception in quiet and noise in postlingually deafened adult CI users. We hypothesized that the IPGE_slope_ and the PLV are two independent measures predictive of speech recognition scores in quiet and in noise, respectively. The hypotheses were partially supported by the results showing that the IPGE_slope_ and the PLV had different relationships with electrode location and age, and that the speech perception scores measured in noise with +5 dB SNR were positively associated with the PLV. However, contrary to our hypothesis, we did not observe significant associations between the IPGE_slope_ and speech perception measured either in quiet or in noise. The result patterns stayed largely the same regardless of fitting methods (window vs. linear) or I/O scales (linear-linear vs. linear logarithmic) used in AGF slope calculation.

### Peripheral Neural Survival and Synchrony

The IPGE_slope_ and the PLV demonstrated different variabilities across electrode locations and age, suggesting that they were measuring different aspects of CN health at least to some extent. These results are aligned with the physiological process of neural degeneration of the CN that occurs in patients with sensorineural hearing loss and are consistent with histological results of human temporal bone studies. Detailed explanations are provided below.

The deterioration of the bipolar SGNs starts at the peripheral axon, which connects the SGN soma to the organ of Corti in the cochlea (Xing et al. 2012). The degeneration of the peripheral axon starts with deterioration of the myelin sheath (Xing et al. 2012) and is followed by axonal dystrophy, segmental swelling (ballooning), and ultimately, loss of the peripheral axon (e.g., Chen et al. 2006; Heshmat et al. 2020; Kumar et al. 2022; Liu et al. 2015; Nadol 1990; Wu et al. 2019; Wu et al. 2023). Even after the complete loss of the peripheral axon, the soma and the central axon of the SGN (i.e., unipolar SGN) can survive for decades (Rask-Andersen et al. 2010), and the proportion of unipolar SGNs increases steadily with age (Wu et al. 2023). This process gives rise to a key difference between acoustic and electric hearing. While the loss of peripheral SGN axons likely contributes to impairment in acoustic hearing (Wu et al. 2019; Wu et al. 2020; Wu et al. 2021), electric hearing can be achieved even without the peripheral axons, as the stimulation can directly reach the somata and/or the central axons of SGNs (Javel & Shepherd 2000). Both axonal dystrophy and demyelination alter many neural properties (e.g., Tasaki 1955; Waxman & Ritchie 1993), which reduce the synchronized discharge of affected CN fibers (e.g., Gonzalez 2019; Heshmat et al. 2020; Kim et al. 2013; Tasaki 1955) as well as across the population of CN fibers (Kandel 2002). Additionally, loss of the peripheral axon and altered membrane properties can shift the action potential initiation site distally to the SGN soma or central axon in electrical hearing (e.g., Hartmann et al. 1984; Javel & Shepherd 2000; Van Den Honert & Stypulkowski 1984), further reducing discharge synchronization across CN fibers (Javel & Shepherd 2000). As explained in He et al. (2024), the PLV is influenced by temporal jitter in spike firing of individual CN fibers as well as discharge synchronization among a population of activated CN fibers across multiple stimulations, which is an ideal tool to assess neural synchrony in the CN in CI patients.

The results of human temporal bone studies demonstrated an age-related loss in both SGN soma and the peripheral axon in adult listeners: Compared with the apical regions, the basal area of the cochlea shows the greatest loss in both SGN soma and peripheral, with peripheral axon showing a substantial greater loss than SGN soma axon (Liu et al., 2015; Makary et al., 2011; Wu et al., 2019; Wu et al., 2023). In one CI case who was 70 years of age at the time of death, a total loss of peripheral axons for all surviving SGNs at the basal half of the cochlea was observed. In contrast, peripheral axon counts in the apical cochlea are similar to those of age-matched, nonimplanted listeners (O’Malley et al. 2024).

Based on the literature reported above, smaller PLVs are expected at more basal electrode locations where substantial degeneration in the peripheral axon occurs in middle-aged and older CI patients, which make up a majority of the patient group tested in this study. Consistent with this expectation, our data showed an electrode location effect on the PLV, with significantly smaller PLVs measured at the basal electrode location than apical locations. Our results did not show the effect of age on the PLV. However, this result does not necessarily conflict with those reported in Wu et al. (2019) and Wu et al. (2023) due to the difference in cases/participants included in these two studies. Specifically, the cases reported in Wu et al. (2019) and Wu et al. (2023) were non-CI listeners with acoustic hearing, while the current study focused on CI users. Further studies using animal models, histological studies in human temporal bones and computational modeling techniques are warranted to determine the crucial factors (e.g., age, surgical trauma, etiology) for neural health of the CN in implanted cases. Finally, the lack of association between the PLV and age calls for further investigations on CI users with a wide age range to determine the contribution of poor neural synchrony in the CN to speech perception deficits in noise experienced by many older CI users.

Unlike the PLV, the IPGE_slope_ is negatively correlated with participant age but not affected by electrode location. The significant correlation between the IPGE_slope_ and age is consistent with the age-related decline in SGN counts in humans reported in different studies (e.g., Liu et al. 2015; Kusunoki et al. 2004; Makary et al. 2011; Wu et al. 2019; Wu et al. 2023) . The lack of difference in the IPGE_slope_ across electrodes is consistent with histological results of some human temporal bone studies reporting similar amounts of SGN loss across cochlear locations (Makary et al., 2011; Wu et al., 2023). However, substantial variations in SGN loss at different cochlear locations have also been reported in the literature (e.g., Hinojosa & Marion 1983; O’Malley et al. 2024). To better understand and interpret our results, we compared the overall trend of IPGE_slope_ across electrode locations with the SGN counts of 26 implanted cases reported in Seyyedi et al. (2013). This temporal bone study was selected for comparison due to the similarities in sample size (28 vs. 26) and etiology of hearing loss (unknown, Meniere’s disease, acoustic neuroma, and trauma), CI devices from the same manufacturer (Cochlear® Ltd.), and the same number of locations/segments (i.e., 4) along the cochlea being investigated. It should be noted that the cases reported in Seyyedi et al. (2013) range from 42 to 92 years of age (mean: 73.46 yrs; *SD*: 12.22 yrs) at the time of death, which is significantly older than the participants tested in our study (Independent sample *t*-test: *t_(52)_* = 3.11, *p* = .003). Figure 7 shows box plots of SGN counts for the implanted ears reported in Tables 5 and 6 in Seyyedi et al. (2013) as a function of cochlear location, along with IPGE_slope_ results measured in our study.

**Figure 7.**
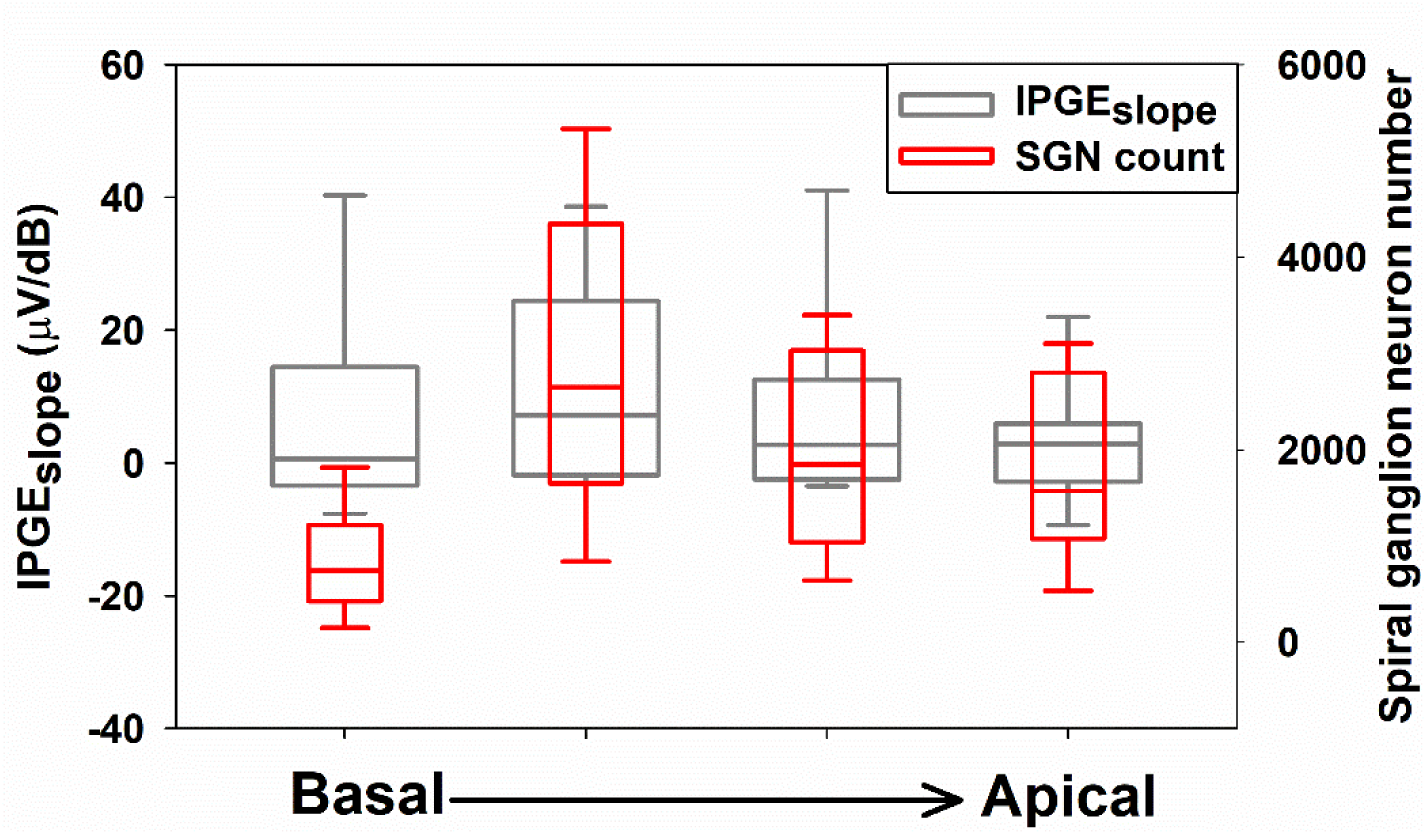
Box plots showing the results of the IPGE_slope_ measured in this study (wide gray boxes) and spiral ganglion neuron counts (narrow red boxes) reported in Seyyedi et al. (2013) across four cochlear regions. Boxes show the range between the first and the third quartile of the data values. The horizontal bars inside the boxes represent the median. The vertical whiskers show the range of values that are within 1.5 interquartile range (IQR) from the boxes. The dots show the 5^th^ and 95^th^ percentile of the data.

Despite the difference in the exact cochlear location where these two measures were taken, the overall trends of these two datasets measured at all except for the most basal location are remarkably similar. For the results measured at the most basal location, SGN count shows a steeper decrease compared to IPGE_slope_ results. Possible factors accounting for this difference include the older age of the cases included in Seyyedi et al. (2013) and the greater degrees of sensorineural hearing loss in these cases due to the stricter CI candidacy criteria used decades ago compared to our study. Both factors result in greater SNG loss at the base of the cochlea than more apical areas. These results, along with those reported in He et al. (2020) and Yuan et al (2022), support the idea that IPGE_slope_ is indicative of SGN survival. Unfortunately, the numeric relationship between IPGE_slope_ and SGN count cannot be determined based on data shown in Figure 7 due to different participants included in our study vs those included in Seyyedi et al. (2013).

It should be noted that both the PLV and the IPGE_slope_ are eCAP-derived measurements.

The eCAP is a near-field recording of the aggregated activity of a group of electrically stimulated CN fibers. As a result, it is affected by firing synchrony both within and across CN fibers as well as the number of activated CN fibers. In addition, each eCAP response used in AGF slope calculation in this study was an average of 50 single sweeps. Its amplitude can be affected by the variability in the peak latencies of the single sweeps, resulting in covariation between IPGE_slope_, an amplitude-based eCAP measurement, and the PLV. Therefore, the trend of positive correlation between the PLV and IPGE_slope_ values observed in this study is not surprising. However, the differences in their associations with electrode location and age indicate that the PLV and the IPGE_slope_ reflect different aspects of CN health. Further studies using computational modeling techniques are warranted to determine the biological underpinnings of these measures and how different types and degrees of neural degeneration of the CN affect these measures.

### Peripheral Neural Synchrony Contributes to Speech Perception in Noise

We observed significant positive associations between the PLV and AzBio scores measured in noise with +5 dB SNR, suggesting that neural synchrony is important for speech perception in high levels of background noise. These results are consistent with and complement the data from our previous study on CI users (He et al. 2024; Skidmore et al. 2023). This observation also agrees with and expands the findings by Harris et al. (2021), where the CAP-derived PLV was shown to be a strong predictor of the perception of time-compressed speech and speech in noise in NH listeners. These results highlight the importance of neural synchrony in the CN in speech perception in listeners with various hearing profiles. The difference between the PLV effects on speech perception across listening environments is likely due to the heightened importance of temporal cues for speech perception in noise (Nie et al. 2006), and the lack of neural synchrony is associated with poor performance in psychophysical tasks requiring fine temporal perception (Zeng et al. 2005). The current results can also provide validation for using the eCAP-derived PLV as a measure of neural synchrony in adult CI users (He et al. 2024). Future studies could measure neural synchrony in the CN in implanted deafened animals using both the eCAP-derived PLV and traditional single-neuron recording methods (e.g., Seki & Eggermont 2003) to further evaluate the validity of using the PLV as an index for neural synchrony in the CN.

### Lack of Significant Associations between the IPGE_slope_ and Speech Perception

The lack of significant associations between the IPGE_slope_ and speech perception in quiet does not support our hypothesis on the contribution of neural survival to speech perception. This result is consistent with those reported by Imsiecke et al. (2021) but not consistent with the observations in a recent study by Zamaninezhad et al. (2023). This discrepancy could be due to some critical differences between the testing materials and methods used in these studies.

Specifically, the German matrix sentences in Zamaninezhad et al. (2023) consisted of syntactically correct but semantically unpredictable sentences, while the AzBio corpus used in the current study and the Hochmair-Schulz-Moser (HSM) sentence test used in Imsiecke et al. (2021) consisted of meaningful sentences on everyday topics; the Freiburg monosyllable test in Zamaninezhad et al. (2023) required the listeners to repeat a single word at a time, while the AzBio and HSM tests required them to repeat a full sentence in each trial. These differences allow the AzBio and HSM sentence tests to better simulate real-life listening situations, but also leave room for the effect of cognitive factors such as working memory (Ingvalson et al. 2015) to modulate the speech perception performance on top of CN health condition. Therefore, it is possible that the association between the IPGE_slope_ and speech perception, if any, has been eclipsed by the individual differences in cognitive factors in the present study. Future research could test both cognitive abilities and the IPGE_slope_ in the same group of CI users to evaluate their relative contributions to speech perception.

In addition, the IPGE_slope_ values in Zamaninezhad et al. (2023) were calculated as the difference between the AGF slopes measured with IPGs of 30 µs and 2.1 µs, but the present study used the AGF slopes between 42 µs and 7 µs for the same calculation. It is possible that the sensitivity of the IPGE_slope_ as an index of CN survival varies with the IPG levels used for eCAP recordings, and further investigation is warranted for optimizing the parameters in the IPGE_slope_ measurement for the purpose of representing CN health condition.

### Frequency Importance Function in Speech Perception Measures

While not a central focus of the present study, in the models evaluating the contributing factors to AzBio scores, we calculated the values of the IPGE_slope_ and the PLV as both unweighted averages across the tested CI electrodes and weighted averages based on the AzBio FIF. The results were similar regardless of whether the AzBio FIF weights were applied, which seemingly contradicts the definition of the FIF (Lee & Mendel 2017). A possible explanation is that the FIF used in the current study was measured in NH listeners (Lee & Mendel 2017), and thus may not be fully generalizable to CI users due to the differences in the weighting of frequency bands in speech perception between CI users and NH listeners (Sladen & Ricketts 2015) and potentially larger individual differences in CI users than NH listeners (Mehr et al. 2001; Bosen & Chatterjee 2016). Furthermore, the weights of adjacent frequencies could differ considerably in some FIFs (Healy et al. 2013), so that estimating the weights of CI electrodes based on a smooth curve fitted to discrete values of the empirically measured AzBio FIF may have limited validity, even if the overall FIF shape of CI users is similar to that of NH listeners. Future research could develop and validate methods for measuring the FIF in CI users and test whether weighting eCAP-derived indices by the CI-based FIF can improve their capability to predict speech perception.

### Methodological Considerations

#### Fitting Method and I/O Scale

One methodological factor to consider when calculating the IPGE_slope_ is the fitting method and I/O scale of the AGF slopes. Skidmore et al. (2022) demonstrated that the window method is broadly applicable to various AGF shapes, and that the window fitting results calculated using linear-linear scale (µV/nC) and linear-logarithmic scale (µV/dB) were highly correlated in human listeners. To further evaluate how the I/O scale affects the calculation of AGF slope at the level of individual electrodes, we compiled the indices of the selected windows (1 to 8, where 1 and 8 correspond to the lower and higher ends of the dynamic range, respectively), and the range of stimulation levels (in CL) corresponding to the selected window of each tested electrode, when calculated both in µV/nC and in µV/dB. When averaged across all electrodes and participants, the selected ranges in CL were similar when using µV/nC (176.1- 186.8 CL) and µV/dB (178.1-188.0 CL), while the selected window index differed by 1 window on average (µV/nC: 5.2, µV/dB: 6.2).

For most of the electrodes, similar window indices and CL ranges were selected when using µV/dB and µV/nC. In particular, for those with a concaved AGF shape similar to the lower half of the S-shaped sigmoid function, a larger window index (typically 7 or 8) was selected when using both µV/dB and µV/nC. However, the inherent differences in the units of dB (re 1 nC) and nC would result in different slope values. When the same or similar windows were selected, the direction of the numerical difference between AGF slopes in µV/dB and µV/nC were dependent on the stimulation level of the selected window. A qualitative observation was that when the stimulation level of the selected window was relatively high, with an upper limit around 175 CL or higher, the slope value calculated in µV/dB was generally larger than µV/nC.

Alternatively, when the stimulation of the selected window was relatively low, with an upper limit around 165 CL or lower, the slope value calculated in µV/dB was generally smaller than µV/nC.

In rare cases, the selected window indices were very different between µV/dB and µV/nC (e.g., for S03 e15 with an IPG of 42 us, windows 8 and 1 were selected when using µV/dB and µV/nC, respectively). Visual inspection of their AGF curves revealed that their AGF was more linearly shaped (rather than concaved) when the amplitude was plotted against CL (a logarithm unit). The linear unit of nC changes at a smaller step when the stimulation changes by 1 CL at lower levels compared to higher levels, while the logarithmic unit of dB (re 1 nC) changes at roughly equal steps across stimulation levels. Such difference would result in larger slopes in µV/nC at lower stimulation levels compared to higher stimulation levels for AGFs with a linear shape when plotted against CL, so that a small window index (corresponding to low stimulation levels) would be selected when using µV/nC.

In short, the numerical difference in AGF slopes between window fitting using µV/dB and µV/nC is influenced by both the shape of the AGF curve and the stimulation range of the electrode. Such subtle numerical differences at the level of individual electrodes may result in different statistical significance in data analysis. However, given the overall similarity in the AGF slopes calculated using the window method in µV/nC and µV/dB, it is unlikely that the general result trends are drastically different across I/O scales when using the window fitting method, as demonstrated by the IPGE_slope_ results in the current study.

#### PLV Analysis Applied to eCAP Recordings

The eCAP PLV analysis methodology used in this study follows that of He et al. (2024), which was adapted from a PLV analysis approach for click-evoked acoustic compound action potentials (CAPs) developed by Harris et al. (2021). The parameters of the analysis need to be adapted because of the difference between the morphologies of eCAPs and click-evoked acoustic CAPs. Schvartz-Leyzac et al. (2025) have recently expressed concerns about the suitability of this adapted PLV method regarding the sampling rate, time window duration, and lack of measurement of a baseline PLV before the stimulation period. However, in human CI users, the shortest possible N1-P2 interpeak latency of the eCAP response is around 0.2 ms (for a review, see He et al. 2017). In a Fast Fourier Transform (FFT) analysis, this corresponds to the shortest possible fundamental period of 0.4 ms or the highest possible fundamental frequency of 2.5 kHz. The sampling rate of Cochlear™ Nucleus® device is 20,492 Hz, and therefore the fundamental frequency and first few harmonics of these shortest possible eCAP waveforms fall below the Nyquist frequency of 10,246 Hz. Thus, this sampling rate, despite being the lowest among all three major CI devices, is sufficient for eCAP recording and is not a factor that could limit the sensitivity of PLV measures. Additionally, in human CI users, the eCAP occurs within the first 1200 μs after stimulus onset (Botros et al. 2007). Therefore, a recording window of 1600 μs is sufficient for capturing the entire eCAP response. It is worth noting that the duration of the time frame used in the FFT analysis, rather than the recording window of the eCAP, affects the frequency range used in the PLV calculation. As demonstrated in He et al. (2024), the number of time frames used to calculate the PLV, when the recording window is fixed at 1600 μs, is not a determining factor for the PLV method. More detailed explanations are in the “Methodological Considerations” section of He et al. (2024).

It is true that the recording window for measuring the eCAP using the neural response telemetry function implemented in the Custom Sound EP software v6.0 (Cochlear Ltd., New South Wales, Australia) does not allow for measurement of a baseline period before the stimulus presentation. However, the duration of the time window can be extended to include 64 samples (around 3,200 μs), which provides an opportunity to compare PLVs calculated for the signal (i.e., eCAP responses) with those calculated for the baseline noise recorded following the eCAP waveform. To fully understand the limitations of the PLV method, we used this extended time window to record the eCAP results in nine participants tested in this study. For the traces measured at each electrode location in these participants, PLVs were calculated for two nonoverlapping 32-sample time windows, consisting of the first and the second half of recorded samples. As has been pointed out previously, the eCAP in human CI users occurs within the first 1200 μs after stimulus onset (Botros et al. 2007), which is fully captured by the first time window. Therefore, the PLV caculated for the first time window was defined as the “signal” PLV, and the PLV calculated for the second time window represent the PLV of the noise floor. The segments of the recorded traces corresponding to the second time window at four electrode locations in the left ear of S06 and the heat maps of time-frequency-specific PLVs are shown in Supplemental Digital Content 1. The segments of these traces corresponding to the first time window and the PLV heat maps are shown in Figure 4. The PLVs measured for the eCAP signal (i.e., the first time window) and the noise floor (i.e., the second time window) at each electrode location measured in these nine participants are shown in Supplemental Digital Content 2. The results of a LMM showed that PLVs calcaulted for the signal were significantly larger than those measured for the noise floor (*F_(1, 52.14)_* = 106.60, *p* < .001), after controlling for electrode location and individual differences.

### Potential Study Limitations

This study has five potential limitations. First, identifying the best parameter and method to quantify the IPG effects is an ongoing research topic. In this study, the IPGE_slope_ was selected to quantify the IPG effect based on the current literature. Future studies may suggest other parameters or quantification methods as preferred. If this is the case, we will reanalyze these data using the preferred parameter and/or method. Second, the PLV and the IPGE_slope_ are eCAP- derived measurements. As a result, the possibility that these measures are affected by non-neural factors cannot be completely excluded. Third, only the association between the PLV and IPGE_slope_ was assessed and reported in this study. The potential associations between the PLV and other parameters that have been used to quantify the IPG effects in published studies are not reported because they are beyond the scope of this study. These associations will be reported in another paper in the near future. Fourth, the present study is that only 28 post-lingually deafened adult participants were included in the study. Most of them were within the age range of 50-85 yrs and had generally good speech perception outcomes. Therefore, the variance in speech perception scores explained by the PLV or the IPGE_slope_ may not represent the variance explained in the entire CI patient population. Further studies in CI users with varied speech perception outcomes are warranted to assess the generalizability of the current observations.

Finally, speech perception was measured only using AzBio sentences, which have high ecological validity but prone to the effects of central auditory processing and cognitive factors. In this study, only one ear was tested for each participant, including those who are bilateral CI users. Therefore, in the current dataset, it is not possible to control for potential individual differences in central auditory processing and cognitive abilities using within-participant between-ear comparisons. Future research can test both ears of bilateral CI users and/or add measurements for central auditory processing and cognitive abilities to pinpoint the crucial factors contributing to speech perception in adult CI users. Finally, the 10-talker babble was used as the competing background noise to assess speech perception performance. It does not fully capture the challenge of understanding speech in more complex environments.

## CONCLUSIONS

The IPGE_slope_ and the PLV are two eCAP-derived indices for different aspects of CN health. The significant positive associations between the PLV and AzBio scores measured in noise with +5 dB SNR suggest that neural synchrony is important for speech perception in challenging listening environments. The lack of association between age and the PLV calls for further investigation to determine whether reduced neural synchrony in the CN is unlikely the primary factor accounting for excessive speech perception deficits in older CI users. Future studies can also investigate the contribution of cognitive factors to speech perception and how they interact with the effects of CN health status, as well as use animal models or computational modeling techniques to better understand the biological underpinnings of the IPGE_slope_ and the PLV.

## Data Availability

Data produced in the present study are available upon reasonable request to the corresponding author.

---

**Supplemental Digital Content 1.**
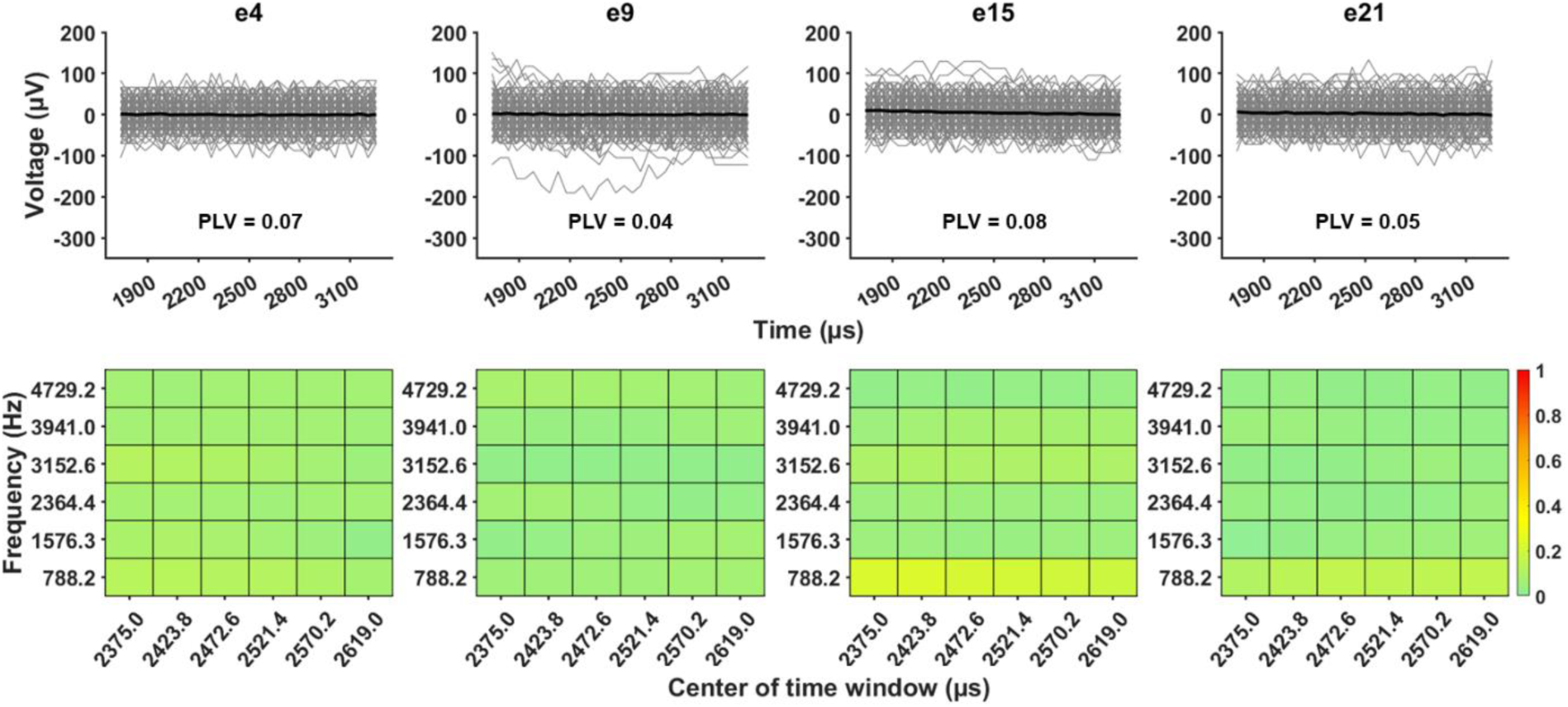

**Supplemental Digital Content 1.**
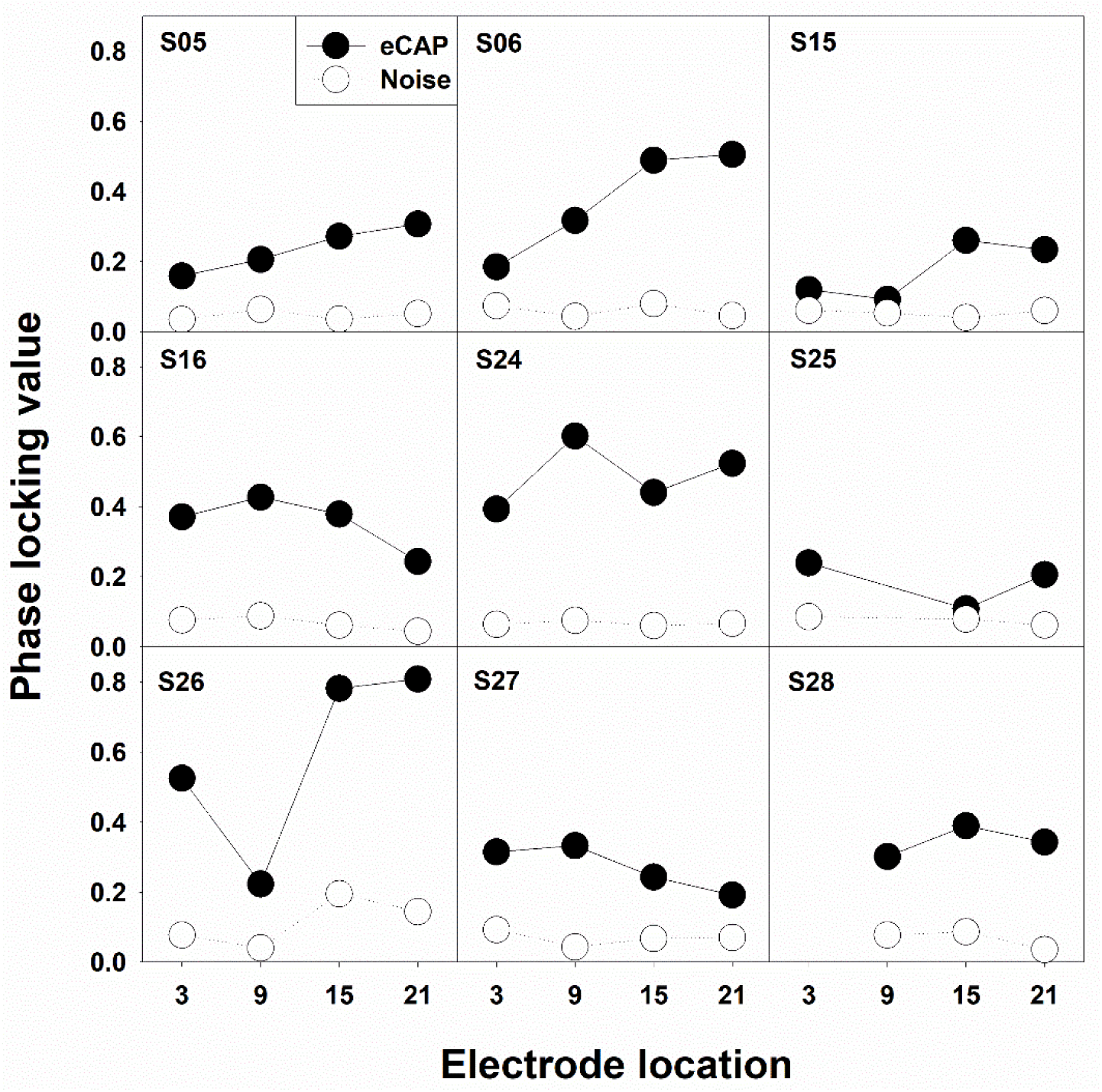

